# Comparison of transmissibility of coronavirus between symptomatic and asymptomatic patients: Reanalysis of the Ningbo Covid-19 data

**DOI:** 10.1101/2020.04.02.20050740

**Authors:** Guosheng Yin, Huaqing Jin

## Abstract

We investigate the transmissibility of coronavirus for symptomatic and asymptomatic patients using the Ningbo Covid-19 data^1^. Through more in-depth and comprehensive statistical analysis, we conclude that there is no difference in the transmission rates of coronavirus between the symptomatic and asymptomatic patients, which is consistent with the original findings in Chen et al.^1^

## 1. Introduction

Since outbreak of the novel coronavirus disease 2019 (Covid-19) in December 2019, the coronavirus has spread over the world at an unprecedented rate. By April 2, 2020, more than 150 countries have been affected by Covid-19 with a total of close to one million confirmed cases and over 47,000 deaths. The Covid-19 has been declared as an international public health emergency by the World Health Organization (WHO).

During the incubation period, a percentage of coronavirus carriers may have no symptoms or minimal symptoms and thus often go undetected. These covert coronavirus carriers may not even be aware of the infection themselves, who however would be confirmed as positive cases if tested using the reverse transcriptase polymerase chain reaction (RT-PCR). If the percentage of asymptomatic carriers is large and if their transmissibility of coronavirus is as high as the symptomatic cases, this would pose a great threat to the public. Therefore, it is critical to determine the percentage and transmissibility of asymptomatic coronavirus carriers in the population.

It has been reported that the viral load detected in the asymptomatic patients was similar to that in the symptomatic patients, which suggests the potential transmissibility of asymptomatic patients.^2^ A familial cluster of five patients in Anyang, China, demonstrated transmission of the coronavirus from an asymptomatic carrier with normal chest computed tomography (CT) who was however tested positive after all the five contacted family members had shown symptoms and confirmed with positive RT-PCR test results.^3^ Another example of coronavirus infection by an asymptomatic patient was a German case through the usual contact in business meetings.^4^

## 2. Issues in the Original Analysis

Chen et al.^1^ carried out an important study using the Covid-19 data from the city Ningbo, China, on the transmission rates of the coronavirus by the symptomatic and asymptomatic cases. The estimated transmission rates for the symptomatic and asymptomatic patients were 0.063 and 0.041 respectively, and the chi-squared test yielded a p-value of 0.288, which indicates that there is no statistically significant difference between the two transmission rates. They further investigated the transmission rates for different relationships and different types of contact with the infected patients including both symptomatic and asymptomatic cases and concluded that the differences in the transmission rates are statistically significant across different relationships and types of contact respectively. The closer the contact with the infected patients, the higher chance of infection.

However, in their original statistical analysis^1^, the chi-squared tests were unduly used because the counts in some cells of the contingency tables are rather small and even zeros, which violates the assumptions of a chi-squared test and thus casts doubt on the validity of the hypothesis test. Moreover, when comparing the transmission rates of symptomatic and asymptomatic cases, Chen et al^4^ included the cases associated with a super-spreader who mainly transmitted the disease in an air-conditioned bus and a buddhism activity gathering. However, this may reduce the generalization of the findings as the super-spreader should be regarded as an outlier and removed from the primary analysis.

## 3. Reanalysis of the Ningbo Covid-19 Data

From January 21st to March 6th 2020, there were 157 symptomatic cases and 30 asymptomatic cases in the Ningbo Covid-19 data^1^. These infected cases resulted in 2147 close contacts with them, among which 2001 exposures were caused by the symptomatic cases and 146 by the asymptomatic cases. The average number of close contacts by the symptomatic cases is 13 and that by the asymptomatic cases is 5, and the difference is statistically significant with a p-value of 6×10^−6^ from a permutation test described in Algorithm 1. The larger number of close contacts by the symptomatic cases may be due to the medical attentions they received after the confirmed diagnosis.

To allow for small cell counts including zeros in the contingency table, Fisher’s exact tests^5^ are used to investigate the difference in the transmission rates between the symptomatic and asymptomatic patients. We consider two scenarios: One combines the numbers of symptomatic and asymptomatic cases as the total number of infected patients, leading to a 2×2 table; and the other separates them, leading to a 2×3 table, as shown in the top panel of Table 1.

**Table 1.** Analysis of transmission rates through close contacts by symptomatic and asymptomatic cases of Covid-19 in Ningbo after removing all the cases associated with the super-spreader

|  | Total close contacts | Infected |  | Uninfected | P-value* |  |
| --- | --- | --- | --- | --- | --- | --- |
|  |  | Symptomatic cases | Asymptomatic cases |  | Combined | Separate |
| Primary analysis of close contacts by symptomatic and asymptomatic cases |  |  |  |  |  |  |
| Symptomatic cases | 1904 (97)<sup>†</sup> | 79 (28) | 15 (4) | 1810 (65) | 0.842 (0.372) | 0.107 (0.076) |
| Asymptomatic cases | 146 | 3 | 3 | 140 |  |  |
| Total | 2050 (97) | 82 (28) | 18 (4) | 1950 (65) |  |  |
| Subgroup analysis by different relationships with infected cases |  |  |  |  |  |  |
| Family | 268 | 37 | 10 | 221 | <10<sup>-6</sup> | <10<sup>-6</sup> |
| Relatives | 400 | 13 | 6 | 381 |  |  |
| Friends | 153 | 23 | 1 | 129 |  |  |
| Coworkers | 57 | 2 | 0 | 55 |  |  |
| Medical | 79 | 0 | 0 | 79 |  |  |
| Others | 1093 | 7 | 1 | 1085 |  |  |
| Total | 2050 | 82 | 18 | 1950 |  |  |
| Subgroup analysis by different types of contact with infected cases |  |  |  |  |  |  |
| Daily activities | 1048 | 69 | 14 | 965 | <10<sup>-6</sup> | <10<sup>-6</sup> |
| Transportation | 167 | 1 | 2 | 164 |  |  |
| Medical contact | 297 | 4 | 0 | 293 |  |  |
| Other contact | 538 | 8 | 2 | 528 |  |  |
| Total | 2050 | 82 | 18 | 1950 |  |  |
| Primary analysis with the estimated rates and 95% confidence intervals |  |  |  |  |  |  |
|  | Odds ratio | Transmission rate of symptomatic cases | Transmission rate of asymptomatic cases | Difference of transmission rates |  |  |
| With super-spreader cases | 1.568 (0.679, 3.620) | 0.063 (0.053, 0.075) | 0.041 (0.017, 0.091) | 0.022 (-0.016, 0.059) |  |  |
| Without super-spreader cases | 1.212 (0.522, 2.815) | 0.049 (0.040, 0.060) | 0.041 (0.017, 0.091) | 0.008 (-0.029, 0.046) |  |  |

\*Combined means p-values are obtained by pooling the numbers of symptomatic and asymptomatic cases together; Separate means p-values are obtained by separating the numbers of symptomatic and asymptomatic cases.
<sup>†</sup>The numbers in the parentheses are associated with the super-spreader; P-values in the parentheses are
obtained when including the cases associated with the super-spreader.

From the results summarized in Table 1, we conclude that there is no significant difference in the transmission rates between the symptomatic and asymptomatic cases, either including or excluding the cases associated with the super-spreader. However, the tests excluding the cases associated with the super-spreader yield larger p-values, 0.842 when combining the numbers of symptomatic and asymptomatic cases and 0.107 when separating them. As a result, there is no evidence in the data to rule out the transmissibility of asymptotic carriers in comparison with symptomatic cases.

The odds ratio, estimated transmission rates and their difference between symptomatic and asymptomatic cases as well as the corresponding 95% confidence intervals are presented in the bottom panel of Table 1. The odds of transmitting the coronavirus to a healthy individual by a symptomatic patient is 1.2 times of that by an asymptomatic patient, which however is not statistically significant as the 95% confidence interval covers one. Furthermore, as the 95% confidence intervals for the difference of transmission rates cover zero, we conclude there is no difference in the transmissibility of coronavirus through close contacts between symptomatic and asymptomatic cases, which is consistent with the findings using Fisher’s exact tests.

The transmission rates under different relationships with the infected cases are significantly different with both p-values <10^−6^ whether combining the symptomatic and asymptomatic cases or not. With regard to different types of contact, the transmission rates are also significantly different with p-values <10^−6^. As expected, the more close contacts with the infected cases, the higher likelihood to contract the coronavirus.

## 4. Conclusion

In summary, we provide a more in-depth analysis of the Ningbo Covid-19 data to examine the difference in the transmissibility of coronavirus for symptomatic and asymptomatic patients. We adopt more appropriate statistical methods, such as Fisher’s exact tests, which require no assumptions on the cell counts in the contingency table. We provide the odds ratio, estimated transmission rates, as well as the corresponding 95% confidence intervals, and the conclusions remain the same that there is no statistically significant difference in the transmissibility of coronavirus between symptomatic and asymptomatic patients.

## Data Availability

The data have been published in a referenced paper.

---

**Figure 1.**
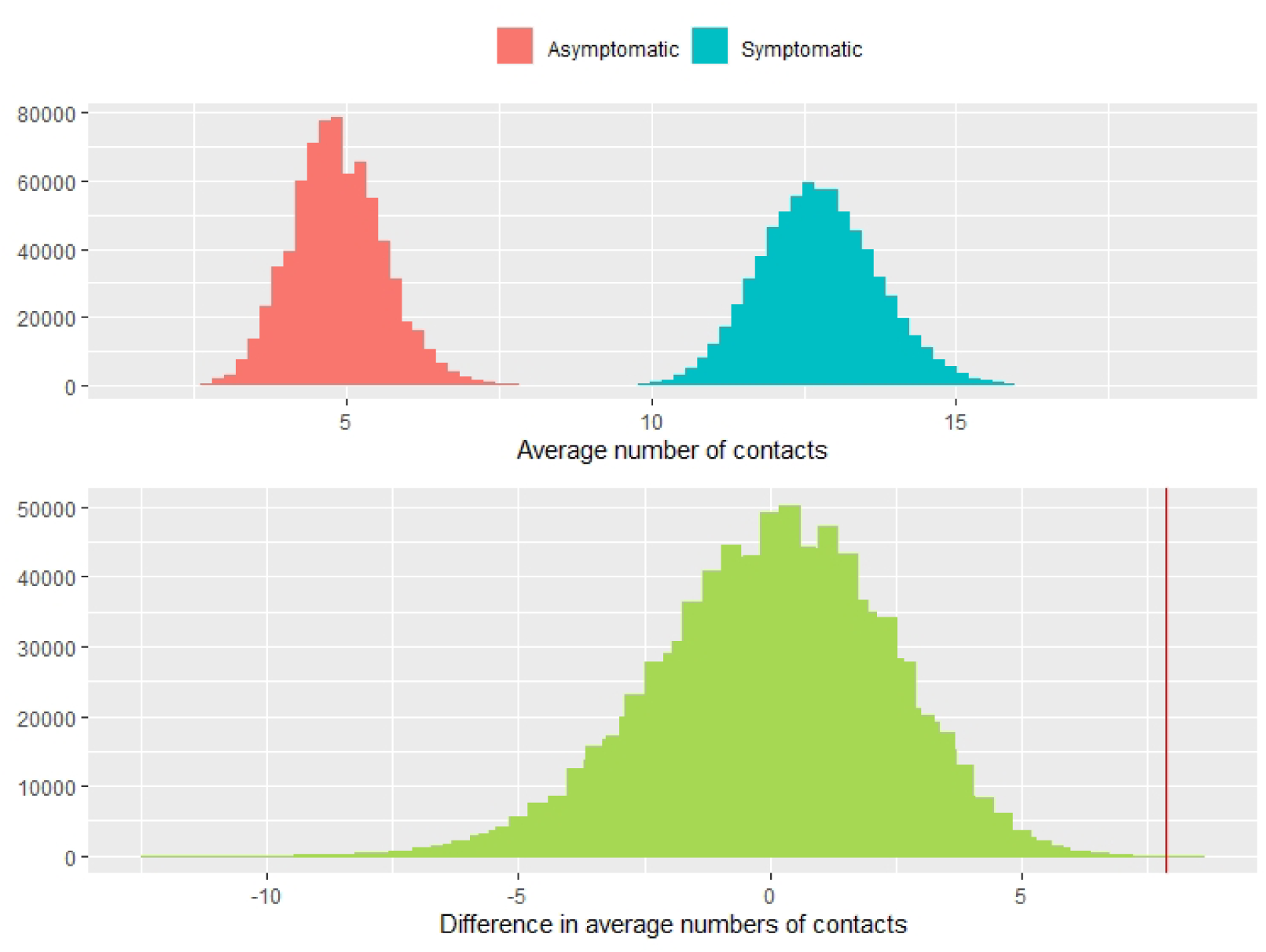
The histograms of average numbers of contacts by symptomatic and asymptomatic cases (top panel) and the difference in average numbers of contacts by the symptomatic and asymptomatic cases in the permutation test (bottom panel). The red vertical line indicates the observed difference in the average numbers of contacts between symptomatic and asymptomatic cases.

**Figure 2.**
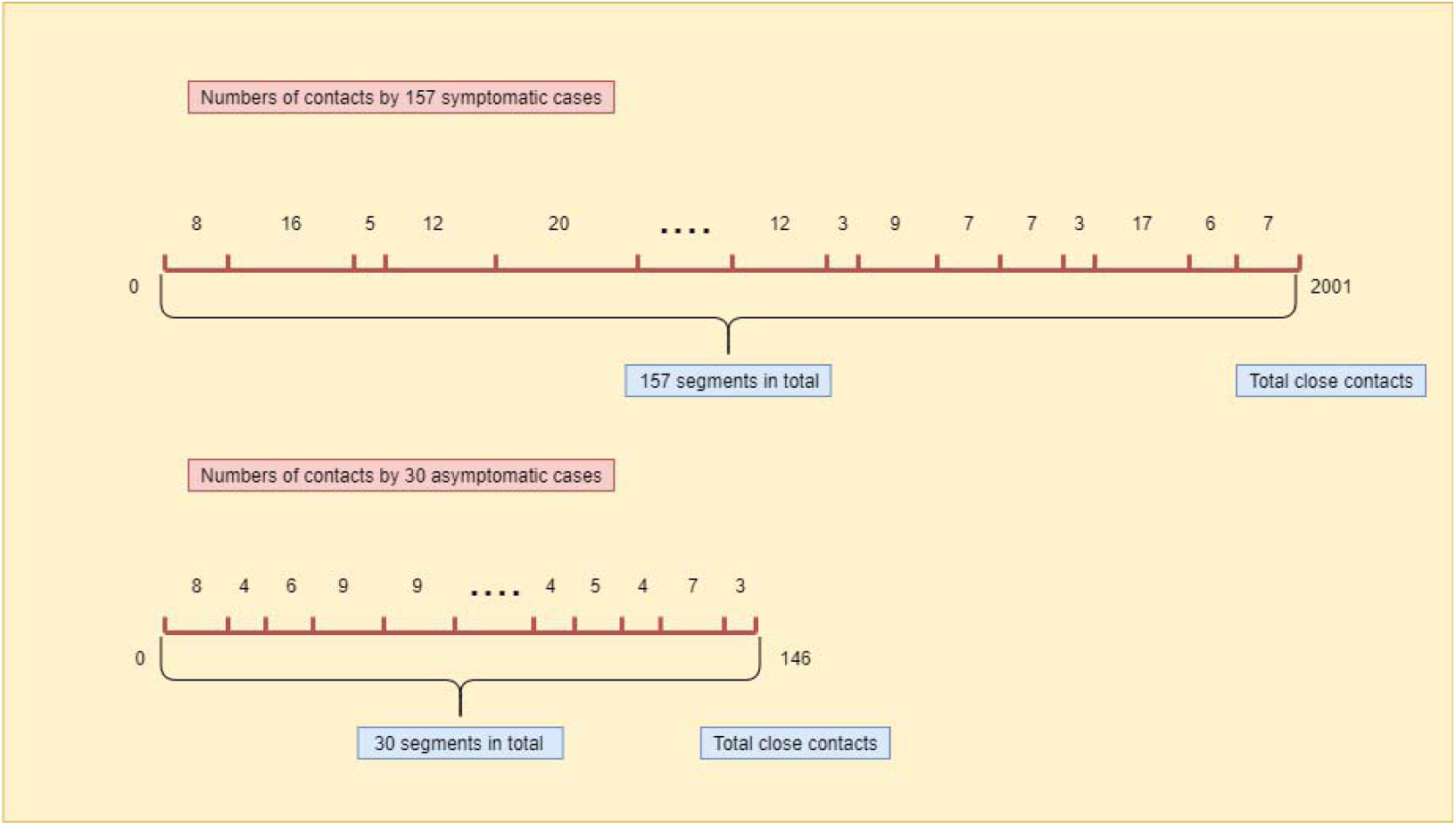
The diagram for the resampling step in the permutation test, where the lengths of segments are randomly generated corresponding to the number of close contacts for each individual patient.

## Algorithm 1. The permutation test

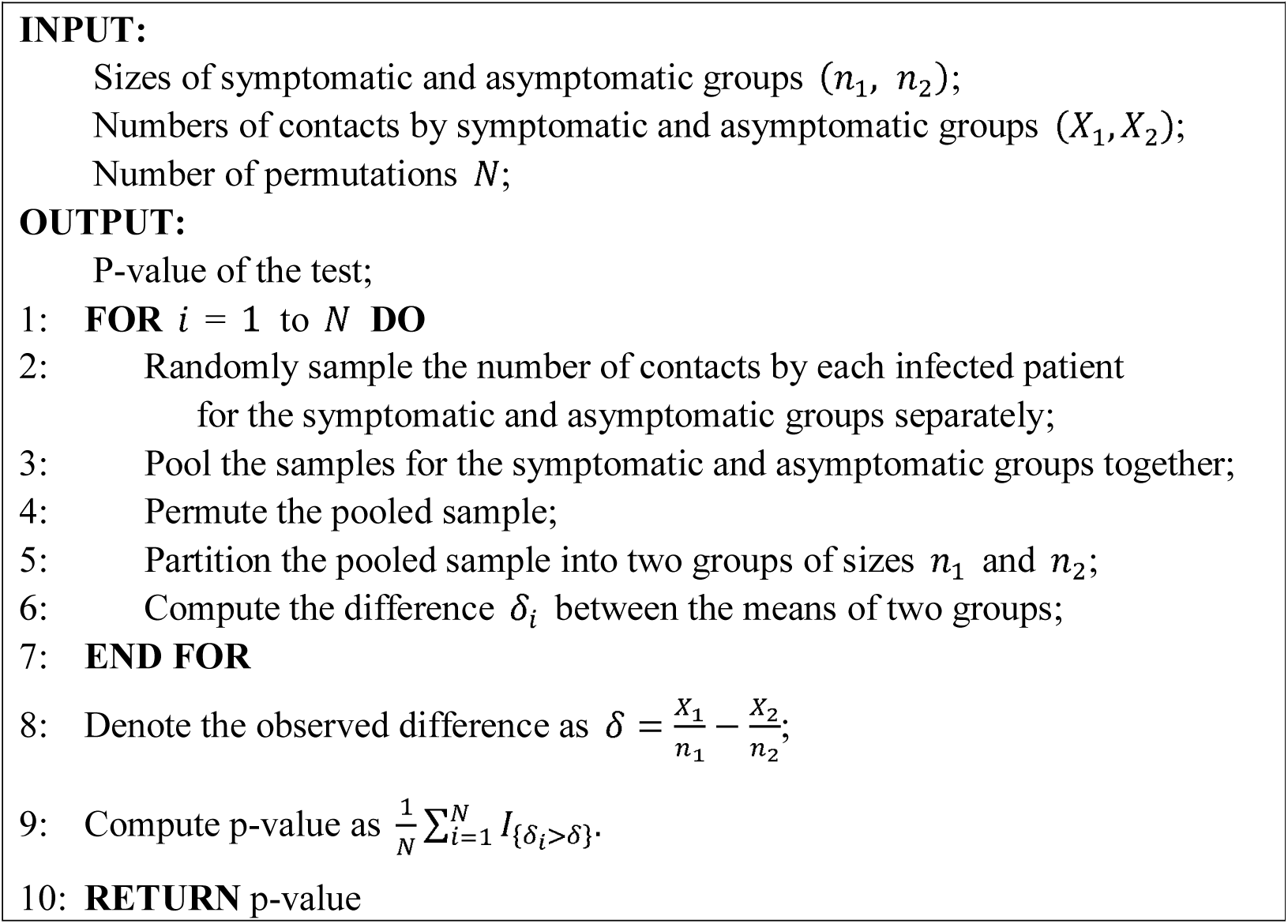

